# SARS-CoV-2 infection is associated with anti-desmoglein 2 autoantibody detection

**DOI:** 10.1101/2022.07.26.22278002

**Authors:** Kerensa E Ward, Lora Steadman, Abid R Karim, Gary M Reynolds, Matthew Pugh, Winnie Chua, Sian E Faustini, Tonny Veenith, Ryan S Thwaites, Peter JM Openshaw, Mark T Drayson, Adrian M Shields, Adam F Cunningham, David C. Wraith, Alex G Richter

## Abstract

Post-acute cardiac sequelae, following SARS-CoV-2 infection, are well recognised as complications of COVID-19. We have previously shown the persistence of autoantibodies against antigens in skin, muscle, and heart in individuals following severe COVID-19; the most common staining on skin tissue displayed an inter-cellular cement pattern consistent with antibodies against desmosomal proteins. Desmosomes play a critical role in maintaining the structural integrity of tissues. For this reason, we analysed desmosomal protein levels and the presence of anti-desmoglein (DSG) 1, 2 and 3 antibodies in acute and convalescent sera from patients with COVID 19 of differing clinical severity. We find increased levels of DSG2 protein in sera from acute COVID patients. Furthermore, we find that DSG2 autoantibody levels are increased significantly in convalescent sera following severe COVID-19 but not in hospitalised patients recovering from influenza infection or healthy controls. Levels of autoantibody in sera from patients with severe COVID-19 were comparable to levels in patients with non-COVID-19-associated cardiac disease, potentially identifying DSG2 autoantibodies as a novel biomarker for cardiac damage. To determine if there was any association between severe COVID-19 and DSG2, we stained post-mortem cardiac tissue from patients who died from COVID-19 infection. This revealed disruption of the intercalated disc between cardiomyocytes that was consistent with separation of the DSG2 protein homodimer. Our results reveal the potential for DSG2 protein and autoimmunity to DSG2 to contribute to unexpected pathologies associated with COVID-19 infection.

## Introduction

Infection with SARS-CoV-2 has been associated with an array of unanticipated symptoms, both during the acute illness and in the immediate and long-term convalescence period. We have previously reported an increased rate of autoantibody detection in individuals post SARS-CoV-2 infection (1). Our previous observation that autoantibodies directed against autoantigens in the skin, skeletal muscle and heart are overrepresented in individuals following severe COVID-19, in comparison to individuals admitted to intensive care for other reasons, suggests a disease-specific effect. Most commonly, skin autoantibodies exhibited an inter-cellular cement pattern by immunofluorescence microscopy, similar to findings in bullous pemphigoid, a blistering autoimmune skin disease due to autoantibodies against desmoglein adhesion molecules (2).

The desmoglein family of proteins play an important role in maintaining tissue integrity through cell-to-cell adhesion within specialized protein complexes called desmosomes. The cadherin family of cellular adhesion molecules includes desmoglein and desmocollin, which through the junctional proteins plakoglobin and plakophilin help secure desmoplakin and keratin filaments to the desmosome structure (3). Desmosomes are found in tissues that experience significant mechanical stress, such as cardiac muscle tissue, gastrointestinal mucosa, and skin epithelia. Desmoglein 1 and 3 are the predominant autoantibody targets for bullous pemphigus and the proteins are strongly expressed in skin; in contrast, desmoglein 2 (DSG2) protein is found in both skin and cardiac tissue. The importance of these proteins is emphasised by the observation that mutations in the desmoglein 2 gene are associated with arrhythmogenic right ventricular cardiomyopathy (ARVC) (4, 5).

Cardiac complications are being increasingly described either during or as post-acute sequelae following SARS-CoV-2 infection (6, 7). These include raised troponin in patients with severe disease (8), viral cardiomyopathy (8) and cardiac pathology on post-mortem hearts (9). Given our previous findings of cardiac muscle autoantibodies (1) we hypothesised that SARS-CoV-2 infection may contribute to cardiac pathology through a direct or indirect autoimmune process.

Here, we have measured desmosomal protein levels and investigated the presence of anti-DSG1, DSG2 and DSG3 antibodies in sera from patients with acute COVID-19 and in those who have recovered from COVID-19 infection. To determine if these autoantibodies are disease specific, we have recruited three comparison cohorts, one with severe influenza infection, a healthy control cohort and a cohort of patients with common underlying cardiac complications. Finally, we examine cardiac tissue, obtained post-mortem from patients who died from COVID-19, to explore the potential clinical relevance of the DSG2 autoantibodies and cardiac damage.

## Methods

### Patient cohorts

We recruited a cohort of thirty-nine patients with severe COVID-19 on the intensive therapy unit (ITU) at the time of sample collection and twenty-five patients that were in convalescence 3-6 months post discharge from the ITU for COVID-19 infection (Ethics: Northwest-Preston Research Committee, ref. 20/NW/0240 IRAS Project ID: 282164). To determine whether there were different rates or patterns of infection following severe or non-hospitalised disease, we included a cohort of forty health care workers from the COvid-19 COnvalescent immunity (COCO) study approved by the London–Camden and Kings Cross Research Ethics Committee (ref. 20/HRA/1817) that had suffered COVID in the first wave in spring 2020 and had tested positive by PCR but had not been hospitalised. As controls for these COVID-19 cohorts we recruited sixteen individuals admitted to the ITU for any reason other than COVID-19. We also recruited two healthy control cohorts; firstly, sera provided by forty-four healthy control subjects prior to the pandemic or by health care workers recruited from the COCO study. The samples from the COCO study were from subjects that were deemed to have never had a COVID-19 infection; they had suffered no clinical symptoms of COVID-19 in the first wave and were negative for IgG, IgA and IgM specific antibodies against the nucleocapsid and spike protein from SARS-CoV-2 – these samples were used to examine levels of circulating desmosomal proteins in sera. Due to insufficient serum volumes being available in the above samples to perform both circulating desmosomal protein and autoantibody assessments we also used sera from a second cohort of fifty COVID-19 naive COCO healthy control sera were used for the desmoglein-2 autoantibody ELISA testing (results shown in figure 1 and table 1)

**Table 1.** Description of clinical cohorts and frequency of positivity for autoantibodies.

|  | **Acute**<br>**COVID**<br>**ITU** | **Convalescent**<br>**COVID ITU** | **Mild COVID**<br>**Convalescent** | **Non-COVID**<br>**ITU** | **Healthy**<br>**Controls** | **Influenza** | **Cardiac**<br>**cohort** |
| --- | --- | --- | --- | --- | --- | --- | --- |
| **Total Individuals,**<br>**n** | 39 | 25 | 40 | 16 | 50 | 42 | 34 |
| **Age (Years),**<br>**median (IQR)** | 54 (50 – 61) | 55 (44.5 – 61) | 44 (35-51) | 57 (46.5 – 68.5) | 44 (37-49) | 42.5 (29-51) | 79 (71-83) |
| **Female, n (%)** | 9 (23) | 5 (20) | 34 (85) | 9 (56) | 34(68) | 20 (47) | 12 (35) |
| **Pre-existing**<br>**cardiac**<br>**Comorbidity, n**<br>**(%)** | 22 (56) | 9 (36) | 4 (10) | 4 (25) | 0 (0) | 6 (13) | 34 (100) |
| **Frequency of positivity for autoantibodies** |  |  |  |  |  |  |  |
| **Skeletal Muscle,**<br>**n (%)** | 9 (23) | 14 (30) | 1 (3) | 0 (0) | n/a | 1 (2) | n/a |
| **Cardiac, n (%)** | 2 (8) | 10 (40) | 0 (0) | 5 (11) | n/a | 3 (6) | 0 (0) |
| **Epidermal, n (%)** | 18 (46) | 13 (52) | 12 (30) | 0 (0) | n/a | 9 (19) | n/a |

**Figure 1:**
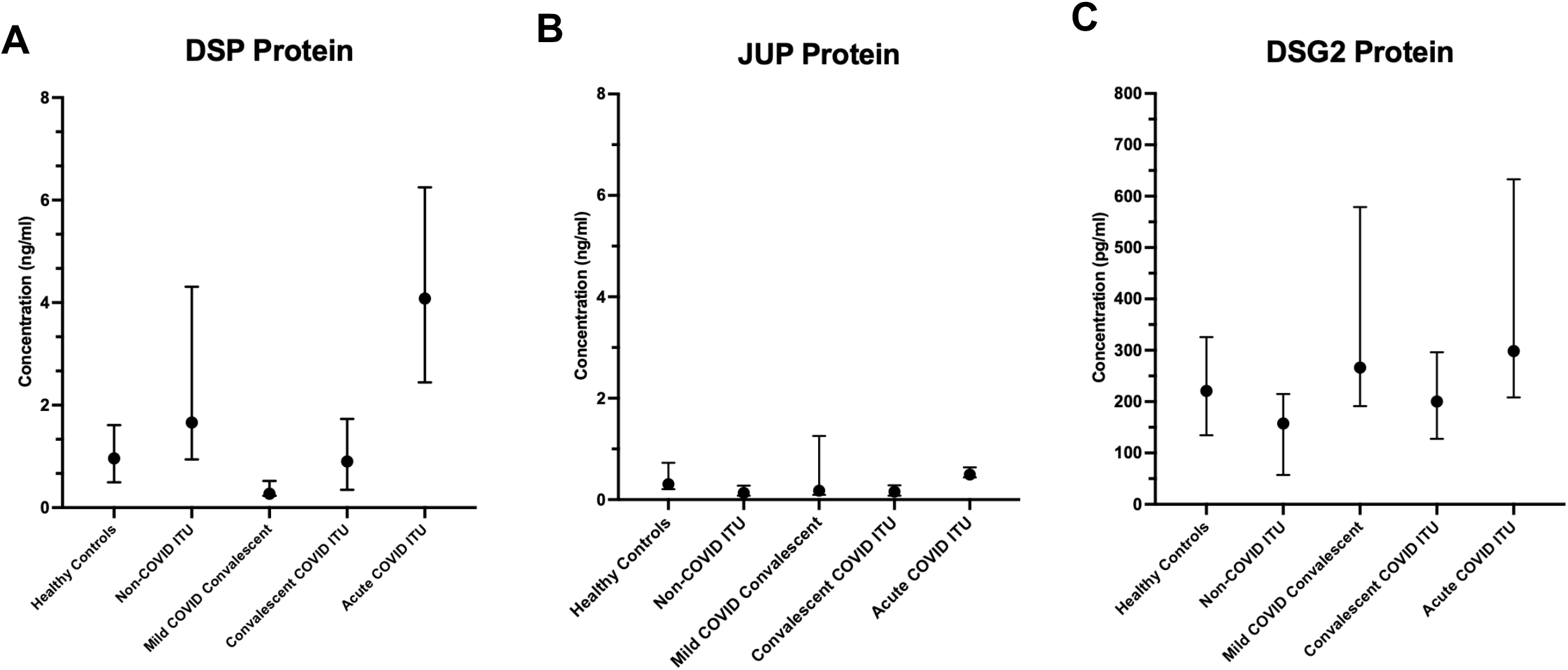
Desmosome proteins detected in serum following SARS-CoV-2 infection. Detection of A) desmoplakin (DSP), B) junction plakoglobin (JUP), and C) desmoglein 2 (DSG 2) protein by ELISA. Results are expressed as median concentration with interquartile range.

To compare results in COVID-19 patients with another group of individuals with severe respiratory viral illness requiring hospitalisation, we obtained acute sera from forty-two individuals who had taken part in the Mechanisms of Severe Acute Influenza Consortium (MOSAIC) study which sampled patients hospitalised with influenza between 2009 and 2011 prior to the COVID-19 pandemic (NHS National Research Ethics Service, Outer West London REC (09/H0709/52, 09/MRE00/67)). To explore whether cardiac comorbidity, aside from ARVC, were associated with DSG2 antibody production we obtained pre-pandemic sera from 34 patients from the Birmingham and Black Country Atrial Fibrillation registry (BBC-AF, IRAS ID 97753, REC reference 12/WM/0344) who had multiple cardiovascular comorbidities; heart failure (*n*=28, 82%), atrial fibrillation (*n*=25, 74%), hypertension (*n*=12, 35%), coronary artery disease (*n*=14, 41%) or myocardial infarction (*n*=9, 26%). Characteristics of each of these cohorts, including previous history of cardiac disease, are described in table 1. Post-mortem tissue, from cases of fatal COVID-19 and pre-pandemic controls without acute cardiac pathology, was collected as part of the LoST-SoCC study (IRAS 193937, Rec: 19/NE/0336). Post-mortems were conducted at the Royal Glamorgan Hospital and Morriston Hospital, South Wales following consent from families. All COVID-19 patients received a positive SARS-CoV-2 swab either in life or at post-mortem, and COVID-19 was given as the cause of death in all cases.

### Indirect Immunofluorescence

Slides with sections of Monkey Heart (REF: S0206), Monkey Skeletal Muscle (REF: S0215), and Monkey Oesophagus (REF: S0205E) (Bio-Diagnostics Ltd, UK) were used to detect anti-cardiac, anti-skeletal muscle, and epithelial autoantibodies respectively. In short, for anti-cardiac antibodies patient sera, positive, and negative controls were each diluted to 1/5 in PBS and 50µl added onto sections. For anti-skeletal muscle and anti-epithelial antibodies the samples were diluted 1/10 with PBS. Slides were incubated at room temperature for 30 minutes before rinsing with PBS and washing for 5 minutes. 30µl of anti-human-FITC-IgG (monkey absorbed) conjugate (REF: J502-5, BioDiagnostics Ltd, UK) was added to each well and incubated at room temperature for 30 minutes. A final PBS rinse and wash was conducted before mounting the slides onto coverslips and reading with a fluorescence microscope with a 495nm exciter filter and 515nm barrier filter.

### ELISAs

#### Commercial ELISAs used to detect serum desmosomal protein concentrations

Human Desmoglein-1 (Invitrogen, Thermo Fisher Scientific, UK; CAT#EH153RB), human Desmoglein-2 (Invitrogen, CAT#EH154RB), human Desmoglein-3 (Invitrogen; CAT#EH155RB), human Junctional plakoglobin (JUP) (Biorbyt; REF: orb563210), and human Desmoplakin (DSP) ELISA kits (Biorbyt; REF: orb564112) were used to detect desmosomal proteins. Serum was run at a dilution of 1/40 as per manufacturers’ instructions. Due to sample availability, not all samples could be run on the protein assays, with the number of samples examined made clear for each result.

### Commercial ELISAs to detect anti-desmoglein-1 and anti-desmoglein-3 autoantibodies

Anti-Desmoglein-1 (Caltag Medsystems, UK; REF: RG-7880EC-D) and anti-Desmoglein-3 (Caltag Medsystems, UK; REF: RG-7885EC-D) autoantibody ELISAs were performed with sera diluted at a dilution of 1/100. Standard protocols as outlined by the product supplier was followed and run using automated machinery (Dynex DSX, Dynex Technologies Limited, UK).

### In-house ELISA to detect anti-desmoglein-2 autoantibody

To detect anti-desmoglein-2 autoantibodies, an in-house ELISA was used. Human desmoglein-2 protein (Biorbyt, UK; REF: orb624074) was diluted to a concentration of 1µg/ml in PBS and 50μl added to each well of a 96-well plate. The assay was incubated in each well overnight at 4°C. The plate was blocked with 200μl 2% bovine serum albumin (BSA) diluted in PBS for one hour, before washing twice with 0.05% PBS-Tween (PBS-T). Each serum was diluted 1/100 with 2% BSA and 100μl of diluted sample was added to each well for one hour at room temperature with gentle shaking. The plate was then washed four times with 0.05% PBS-T. Anti-human-IgG-HRP conjugate (University of Birmingham, UK) was diluted to 1/12,000 in 2% BSA and 100μl was added to each well before the plate was incubated for 30 minutes at room temperature with gentle shaking. The plate was then washed four times with 0.05% PBS-T before the addition of 100μl of 3,3’,5,5’-Tetramethylbenzidine (TMB; Bio-Rad Laboratories Inc., USA; REF: BUF056B) which was incubated for 20 minutes at room temperature with gentle shaking. Finally, 100μl of 0.2M sulphuric acid was added to each well and the plate was read using a microplate reader with a 450nm filter and the results expressed as the optical density (OD). The protocol was run using automated machinery (Dynex DSX, Dynex Technologies Limited, UK).

### Immunohistochemistry to examine DSG2 staining patterns on post-mortem cardiac tissue

Immunohistochemistry was undertaken on cardiac tissue from three post-mortem samples from individuals who died from non-COVID-19 causes and eight individuals who died from COVID-19. Lung tissue from the COVID-19 patients was also examined to rule out non-specific staining. Samples were stained using a mouse anti-DSG2 antibody (AH12.2: sc-80663, Santa Cruz Biotechnology, Texas, USA) by standard IHC techniques. Formalin fixed, paraffin embedded samples were deparaffinised and rehydrated and following low temperature retrieval (ALTER) (10), immunostained on a Dako Autostainer. Anti-DSG2 antibody was applied for 1 hour at 2μg/ml and visualised with Vector Excel mouse kit and ImmPACT NovaRED (2B Scientific).

### Statistical Analysis

All results are recorded as median with interquartile range (IQR) where *n* represents numbers of individual samples. Comparisons between two groups were made using the Mann Whitney U test. Multiple comparisons were made using the Kruskal-Wallis test with Dunnet’s multiple comparisons post-hoc test.

## Results

### Increased serum DSG2 protein levels selectively associate with acute, severe COVID-19

Desmoplakin (DSP) protein was detectable and at higher levels in both acute COVID-19 ITU patients (86% [n=18/21], 4.08ng/ml [2.45-6.25]) and individuals admitted to the ITU for reasons other than COVID-19 infection (68% [n=13/19], 1.66ng/ml [0.94-4.31]) compared with healthy controls (42% [n=13/33], 0.95ng/ml [0.50-1.61], p=0.0004) (**Figure 1A**) Junction plakoglobin (JUP) protein levels found no discernible pattern that discriminated COVID-19 infection or acute disease across the cohorts (**Figure 1B**) along with DSG1 and DSG3 protein levels (data not shown).

DSG2 protein was most likely to be detected, and at the highest concentration, in individuals from the acute COVID-19 ITU group (**Figure 1C**), compared to individuals admitted to the ITU for reasons other than COVID-19 infection and healthy controls (48% [n=10/21], 298pg/ml [208-633] for the COVID-19 ITU group and 16% [n=13/19], 157pg/ml [57.2-215], *p*=0.0007 for non-COVID-19 ITU group and 21% [n=7/33], 222pg/ml [140-332], *p*=0.0068 for healthy controls). This suggests a potential relationship between DSG2 and SARS-CoV-2 infection in more severe COVID-19 presentations.

### Indirect immunofluorescent staining was more likely to be associated with anti-DSG2 rather than anti-DSG1 or 3 autoantibodies

We found an increased presence of skeletal, cardiac and epidermal autoantibodies in the acute COVID ITU group, with 23%, 8%, and 46% patients positive respectively. These high levels were also observed in convalescent COVID ITU patients where positivity increased to 30% skeletal, 40% cardiac, and 52% epidermal autoantibodies. In comparison, cardiac autoantibodies were the only autoantibodies detected in 6% non-COVID ITU patients with low levels also observed in patients with influenza. The cardiac samples had no evidence of anti-cardiac antibodies to suggest their disease or DSG2 level might be related to ARVC. As inter-cellular cement skin autoantibodies are associated with anti-DSG1, 2 and 3 autoantibodies, we wanted to determine which DSG autoantibodies the immunofluorescence findings were associated with. We tested the 30 samples from the COVID groups that were positive for skin autoantibodies (inter-cellular cement staining) by immunofluorescence and found none of these samples were positive for anti-DSG1 or anti-DSG3 autoantibodies. In contrast, we found 80% (n=24/30) had DSG2 autoantibody levels higher than the top interquartile range seen in the healthy control cohort (median OD=0.35 [0.22-0.48], **Figure 2**)

**Figure 2:**
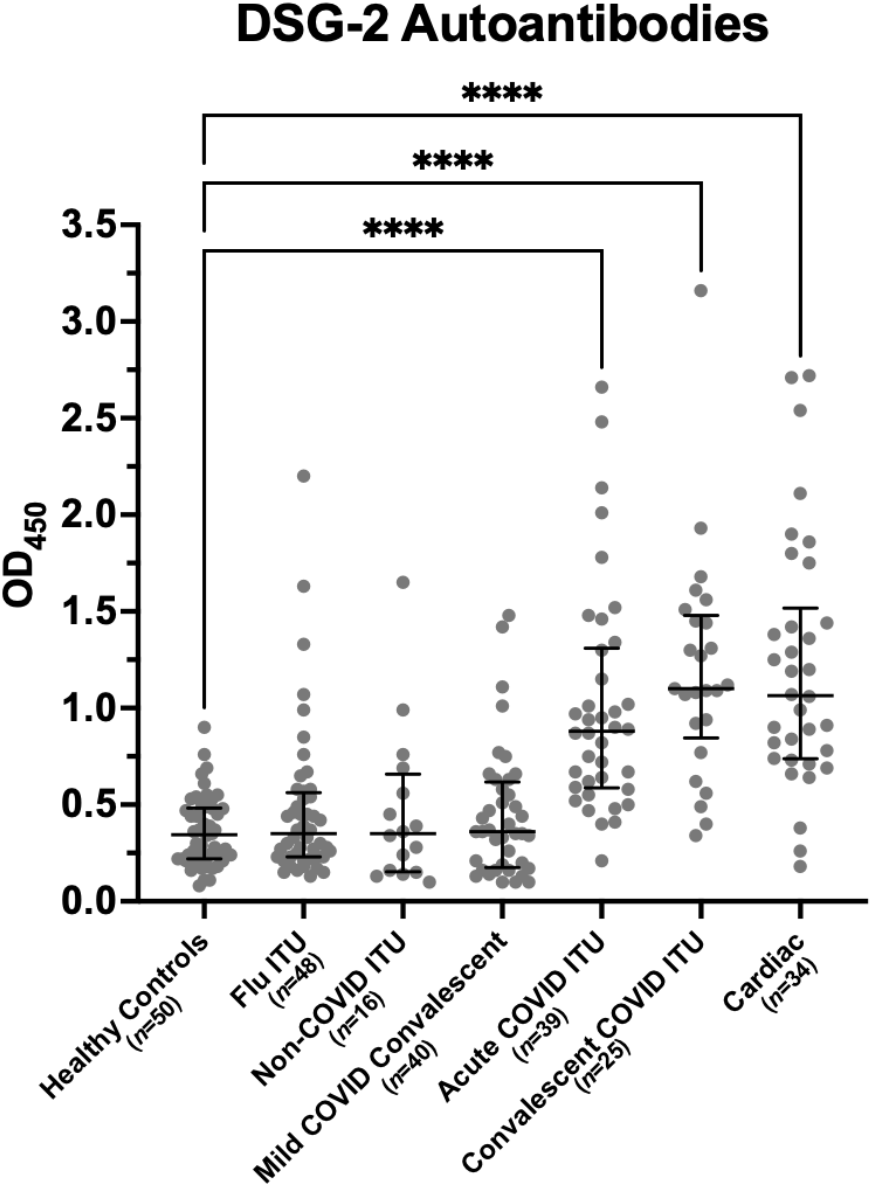
Desmoglein 2 autoantibody concentrations are high in patients post severe SARS-CoV-2 infection. Detection of desmoglein 2 autoantibodies in COVID-19 cohorts by ELISA; convalescent COVID Intensive therapy unit (ITU), acute COVID ITU, and mild COVID convalescent, compared with non-COVID control cohorts; non-COVID ITU, Influenza (flu) ITU, and healthy controls (N=252, p<0.0001). The flu ITU, non-COVID ITU, and mild COVID convalescent cohorts were all not statistically significant compared to healthy controls. Results are expressed as median OD (optical density) with interquartile range. Figure in brackets indicates number of sera per group. **** indicates a significant difference between groups at a p value of <0.0001.

### DSG2 autoantibodies are detected selectively in patients with severe COVID-19

To investigate whether the observed increase in DSG2 protein was associated with DSG2 autoantibody production, we developed an in-house ELISA to detect DSG2 IgG autoantibodies. Levels of DSG2 IgG autoantibodies were significantly elevated in sera from both acute (median OD=0.90[0.60-1.26], n=39, *p*<0.0001) and convalescent (median OD=1.10[0.85-1.48], n=25, *p*<0.0001) COVID cohorts following severe disease when compared with healthy controls (median OD=0.35 [0.22-0.48] n=50) (**Figure 2**). Sera from the cohort of convalescent individuals following mild disease did not have elevated DSG2 autoantibody levels (median OD=0.37 [0.19-0.58], n=40, *p*>0.9999), nor did sera from a non-COVID cohort of ITU patients (median OD=0.35 [0.16-0.71], n=16, *p*>0.9999), nor sera from a cohort of patients in convalescence after influenza infection (median OD=0.35 [0.23-0.56], n=48, *p*>0.9999).

### DSG2 autoantibodies are increased in patients with underlying cardiac comorbidities

Due to the increased rate of autoantibodies against cardiac tissue in our COVID-19 cohorts, we assessed levels of DSG2 autoantibodies in sera from patients with underlying cardiovascular conditions. These studies found antibody levels similar to those found in the COVID-19 patients with severe disease (median OD=1.07 [0.74-1.44], *n*=34, *p*<0.0001) (**Figure 2**). The type of cardiac comorbidity, nor severity of disease as stratified by ejection fraction, Troponin T, NT-proBNP, IL-6 nor hsCRP, did not predict the presence of DSG2 autoantibodies. We obtained history on previous cardiac comorbidity in each of the groups examined (**Table 1**), but found no relationship with the raised DSG2 autoantibody levels observed in the severe COVID-19 patients in both the acute and convalescent setting.

### Intercalated disc staining for DSG2 is abnormal in COVID-19 post-mortem cardiac tissues

All hearts from individuals who did (n=8) or did not (n=3) die from COVID-19 causes (n=3) showed variable weak to strong staining of DSG2 corresponding to the site of the intercalated discs (**Figure 3**). Areas were also observed with no staining due to post-mortem degradation of the tissues. In the non-COVID-19 post-mortem heart tissues DSG2 staining consistently presented as a single, distinct band (**Figure 3A**). In the post-mortem heart tissues from COVID-19 positive patients (n=8) also showed a similar staining pattern, but in addition there were distinct areas of double-banded staining observed, which are indicative of the separation of the intercalated disc (11) (**Figure 3B**). Importantly, this distinct ‘double’ banding pattern was not seen in non-COVID cardiac muscle, even when there was discontinuation of the muscle as a result of post-mortem degradation. No DSG2 staining was observed in the post-mortem lung tissue (n=8) examined from COVID-19 patients.

**Figure 3:**
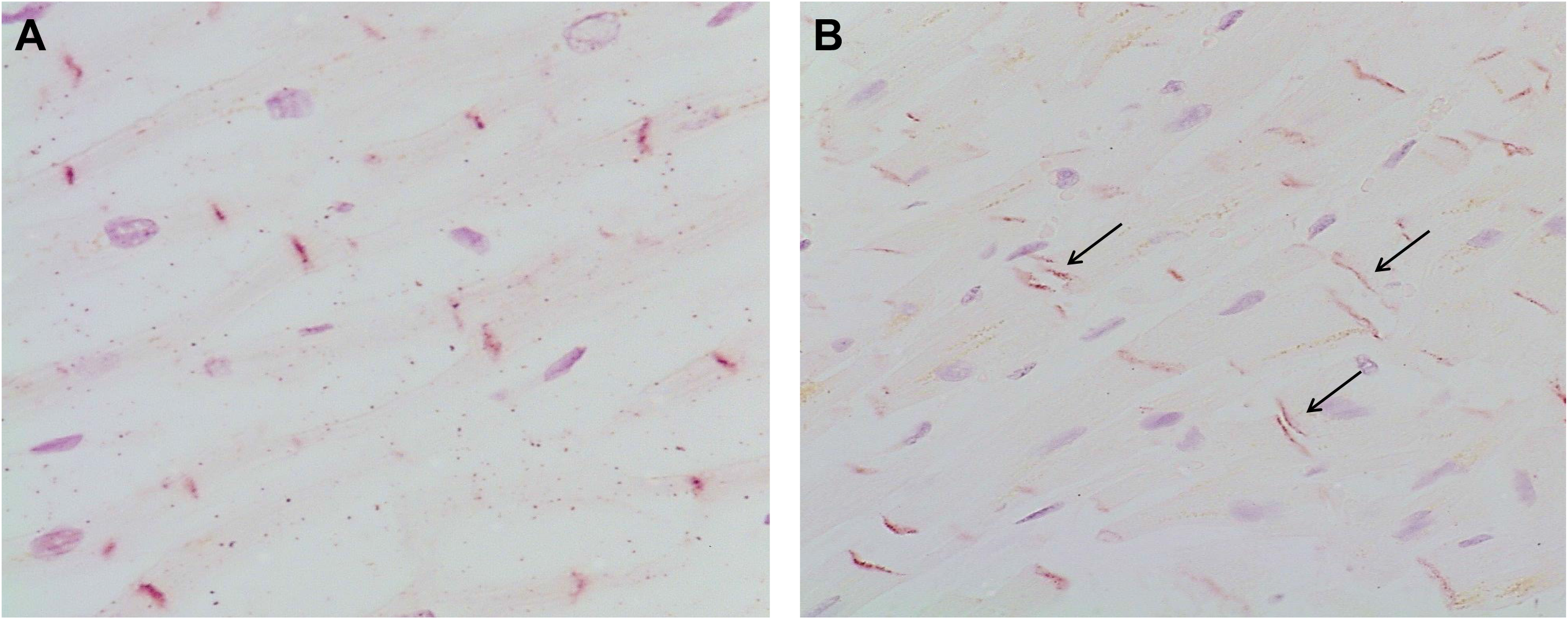
Desmoglein 2 immunohistochemical staining of post-mortem cardiac tissue. Non-COVID post-mortem heart tissue (A) shows uniform single banded staining for DSG2 (red) corresponding to the site of intercalated discs. Cardiac muscle from COVID post-mortem cases (B) showed single and areas of notable double banded staining for DSG2 (arrowed). Images representative of 3 hearts from non-COVID-19 patients and 8 patients who died from COVID-19.

## Discussion

In this study, we have found raised DSG2 protein and a higher frequency of DSG2 autoantibody production in patients with or following severe COVID-19. Increased anti-DSG2 autoantibodies are not induced to all infections as evidenced by the relative absence of anti-DSG2 responses in other groups including a cohort of influenza patients. Furthermore, the detection of perturbed DSG2 staining in post-mortem human heart tissue from COVID-19 patients reveals structural changes of the cardiomyocyte intercalated discs which are rich in DSG2.

There is an extensive literature on the association between numerous viral infections and autoimmunity. This can be due to a number of mechanisms including molecular mimicry, epitope spreading and bystander activation. COVID-19 is no exception with clinical syndromes such as autoimmune cytopenia, Guillain-Barre syndrome, autoimmune encephalitis, lupus and acquired haemophilia being reported (12). COVID-19 infection results in the post-translational modification of a vast number of proteins with high intrinsic propensity to become autoantigens, offering an explanation for the diverse autoimmune complications observed (13).

Although low-titre, transient autoantibodies occur with many acute viral infections (14) their clinical significance and pathogenic potential are uncertain. We have previously reported an increase in autoantibodies post-COVID-19 (1) with a specific pattern of autoantibodies against cardiac, skin and muscle and shown that these were still present at 6 months post-infection. Similarly, in this study we find an increased rate of DSG2 autoantibodies and that these persist to 6 months following an episode of severe COVID-19. The finding of persistent autoantibodies raises the possibility that there may be an ongoing clinical consequence; however, future study is required of longitudinal samples with clinical follow up data to establish causation.

DSG2 autoantibodies have the potential to be pathogenic and have been associated with arrhythmogenic right ventricular cardiomyopathy (ARVC) and familial dilated cardiomyopathy (15). Furthermore, DSG2 autoantibodies have been shown to disrupt DSG2 protein function with the consequence of interfering with cell-to-cell adhesion (15, 16). Consequent intercellular space widening at the level of the intercalated disc (desmosomes/adherens junction) and reduction in action potential velocity results in the increased arrhythmia susceptibility observed (17).

Cardiac injury following COVID-19 infection is now well described in the literature, although the mechanisms behind this remain uncertain (7). Clinical studies have reported post-covid complications such as thromboembolism, ischaemia, arrhythmias, conduction defections and myocarditis (18-20) and post-mortem studies have found increased neutrophil extracellular trap (NET) formation and mononuclear cell infiltration (9). Given that cardiac involvement is an important and long-term consequence of COVID-19, and potential also within the spectrum of syndromes observed in “long-COVID”, we examined post-mortem heart tissue for DSG2 protein. We found that DSG2 is localised to the intercalated discs, confirming previous studies, and importantly that these discs were only found to be widened in COVID-19 tissue samples as is seen in ARVC (4, 17). Previous in vitro studies for ARVC have found that inhibition of DSG2 binding, or mutation of DSG2 protein, disrupts intercalated discs and subsequently the cell to cell contact necessary for cardiomyocyte adhesion. It is increasingly recognised that DSG2 is a multifunctional protein and may have a role in carcinogenesis (21), angiogenesis and re-localisation of actin (22), early haematopoietic development in the bone marrow, in particular myeloid progenitors (22), and intestinal epithelial apoptosis and haemostasis (23). Future studies will have to examine how DSG2 autoantibodies may interfere with DSG2 protein function and the clinical consequences of this, but our findings allow us to propose that DSG2 autoantibodies are disrupting intercalated discs and therefore normal cardiac function. If replicated in larger studies with clinically-associated data, this may offer a potential biomarkers for the multiple and diverse long term sequelae following COVID-19 infection.

In conclusion, the findings of DSG2 autoantibodies and also intercalated disc widening offer a potential autoimmune mechanism for some of the complications found post-COVID-19 infection. Further studies in well characterised clinical cohorts, with comorbid complications, are required to prove whether the presence of DSG2 antibodies serves as a marker of post-acute COVID-19 syndrome or long COVID. Detailed mechanistic studies are required to reveal the link between autoimmunity to DSG2 and the long-term sequelae of SARS-CoV-2 infection.

## Data Availability

All data produced are available upon request to the authors.

## Acknowledgements and funding

The authors would like to acknowledge the staff of the University of Birmingham Clinical Immunology Service for facilitating laboratory studies. The convalescent health care worker study (COCO) was carried out at the National Institute for Health Research (NIHR)/Wellcome Trust Birmingham Clinical Research Facility.

This study was funded as part of the UK Coronavirus Immunology Consortium funded by NIHR and UKRI. This paper presents independent research supported by the NIHR Birmingham Biomedical Research Centre at the University Hospitals Birmingham NHS Foundation Trust and the University of Birmingham.

Post-mortems were performed by Dr Esther Youd, Royal Glamorgan Hospital, Cwm Taf University Health Board and Dr Gareth Leopold, Morriston Hospital, Swansea Bay University Health Board. Post-mortem sample collection was funded by an MRC Clinical Research Training Fellowship and the Welsh Assembly Government

